# Development and Validation of Sex-Specific Hip Fracture Prediction Models using Electronic Health Records

**DOI:** 10.1101/2022.10.26.22281584

**Authors:** Gloria Hoi-Yee Li, Ching-Lung Cheung, Kathryn Choon-Beng Tan, Annie Wai-Chee Kung, Timothy Chi-Yui Kwok, Wallis Cheuk-Yin Lau, Janus Siu-Him Wong, Warrington W.Q. Hsu, Christian Fang, Ian Chi-Kei Wong

**Author notes:** **Correspondence and reprint requests:** Ching-Lung Cheung, PhD, Department of Pharmacology and Pharmacy, The University of Hong Kong, Pokfulam, HONG KONG.

## Abstract

**Background:** Hip fracture is associated with immobility, morbidity, mortality, and high medical cost. Due to limited availability of dual-energy X-ray absorptiometry (DXA), hip fracture prediction models without using bone mineral density (BMD) data are essential. We aimed to develop and validate 10-year sex-specific hip fracture prediction models using electronic health records (EHR) without BMD.

**Methods:** In this population-based study, the derivation cohort comprised 161,051 public healthcare service users (91,926 female; 69,125 male) in Hong Kong aged≥60. Sex-stratified derivation cohort was randomly split to 80% training and 20% internal testing datasets. An external validation cohort comprised 3,046 community-dwelling participants. With 395 potential predictors (age, diagnosis and drug prescription records from EHR), 10-year sex-specific hip fracture prediction models were developed using stepwise selection by logistic regression (LR) and four machine learning (ML) algorithms (gradient boosting machine, random forest, eXtreme gradient boosting, and single-layer neural networks) in the training cohort. Model performance was evaluated in both internal and external validation cohorts.

**Findings:** In female, the LR model had the highest AUC (0.815) and adequate calibration in internal validation. Reclassification metrics showed ML algorithms could not further improve the performance of the LR model. Similar performance was attained by the LR model in external validation, with high AUC (0.841) comparable to other ML algorithms. In internal validation for male, LR model had high AUC (0.818) and it outperformed all ML models as indicated by reclassification metrics, with adequate calibration. In external validation, the LR model had high AUC (0.898) comparable to ML algorithms. Reclassification metrics demonstrated that LR model had the best discrimination performance.

**Interpretation:** Even without using BMD data, the 10-year hip fracture prediction models developed by conventional LR had better discrimination performance than the models developed by ML algorithms. Upon further validation in independent cohorts, the LR models could be integrated into the routine clinical workflow, aiding the identification of people at high risk for DXA scan.

**Funding:** This study was funded by the Health and Medical Research Fund, Food and Health Bureau, Hong Kong SAR Government (reference: 17181381).

## Introduction

Osteoporosis is a prevalent disease characterized by low bone mass and deterioration in bone strength and microarchitecture, which leads to increased risk of fragility fracture. Among all fragility fractures, hip fracture is known to be associated with high immobility, morbidity, and mortality. Earlier projection in 1990s demonstrated that there will be around 4.5-6.26 million hip fractures globally by 2050, with half of them from Asia.^1,2^ This concurs with our recent projection that the number of hip fracture in Asia will reach 2.56 million in 2050, leading to an annual direct medical cost of around USD15 billion in Asia.^3^ Given that hip fracture is associated with high medical cost, prevention of hip fracture is not only essential for people at high risk and their caregivers, but also the healthcare system and society.

Dual-energy X-ray absorptiometry (DXA) is the gold standard for measurement of bone mineral density (BMD) and diagnosis of osteoporosis. It is also an important facility to predict fracture. Yet, its availability is considerably low, especially in the developing countries and regions.^4^ Even a majority of European countries had insufficient provision of DXA machines for the general population to meet the requirements of practice guidelines.^5^ The average waiting time for DXA scan in European countries could be as long as 180 days.^5^ Due to the limited resources for DXA scan services, it is important to develop a fracture risk prediction model without BMD data as a routine screening tool in public healthcare setting, which facilitates the prioritization of people at high risk for DXA scan, aiding early diagnosis and timely treatment of osteoporosis.

Existing prediction tools, such as FRAX, were developed using data mainly from Caucasians.^6^ We previously found that ethnic-specific clinical risk factors outperformed the performance of FRAX in Hong Kong,^7^ demonstrating the importance of developing a population-specific hip fracture prediction tool. Recently, machine learning (ML) algorithms were applied to develop fracture risk prediction models.^8-10^ Notably, most ML models were developed among people in Europe and United States, mainly used to predict the short-term fracture risk in up to five years.^8-10^ In this study, we aimed to develop and validate models that predict the 10-year risk of hip fracture for individuals in Hong Kong using age, diagnosis and drug prescription data in the form of electronic health records (EHR), but in the absence of conventional clinical parameters such as BMD, height, weight and body mass index (BMI). To account for sex-specific factors contributing to the different causes of osteoporosis and hip fracture incidence between the two sexes, these prediction models were separately developed and validated in female and male.

## Materials and Methods

### Data Source

Anonymized medical records were retrieved from the Clinical Data Analysis and Reporting System (CDARS), a large and representative electronic medical database in Hong Kong managed by the Hong Kong Hospital Authority (HA). The HA is a public healthcare service provider that manages 43 hospitals and institutions, and 122 outpatient clinics, serving >80% of hospital admissions. Approximately 98% of hip fracture in Hong Kong was admitted to HA hospitals,^11^ and the hip fracture coding in CDARS was previously validated with a positive predictive value (PPV) of 100%,^12^ suggesting that CDARS data is representative and accurate, particularly for hip fracture. The medical records available in CDARS comprise demographics, prescription (British National Formulary [BNF]), diagnosis (International Classification of Disease, 9^th^ revision, Clinical Modification [ICD-9-CM]), admission, procedures, and laboratory tests.

### Study design and cohort

Figure 1 illustrates the study design. As of 31 December 2005 (index date), about 740,000 public healthcare service users aged ≥60 had admission records at in-patient, out-patient, or accident & emergency services from 1 January to 31 December 2005 in CDARS. Approximately one-third of them were randomly selected. Individuals with complete follow-up from 1 January 2006 till the study end date on 31 December 2015 were included in the derivation cohort. The outcome of interest was the 10-year risk of developing hip fracture, which was identified by ICD-9-CM code of 820.xx.^12^ The derivation cohort was sex-stratified, and each sex-specific sub-cohort was randomly split into the training (80%) and internal testing (20%) datasets. Conventional statistical model and ML algorithms were used to develop the prediction models in the training dataset, followed by validation in the internal testing dataset. Performance of the prediction models were further assessed in the external validation cohort comprising participants aged ≥60 from the Hong Kong Osteoporosis Study (HKOS), which was described elsewhere.^13^ Briefly, the HKOS comprised >9,000 community-dwelling Southern Chinese participants, who were followed using EHR from CDARS. The external validation cohort comprised 3,048 HKOS participants aged ≥60 as of 31 December 2005, without overlap with the derivation cohort.

**Figure 1.**
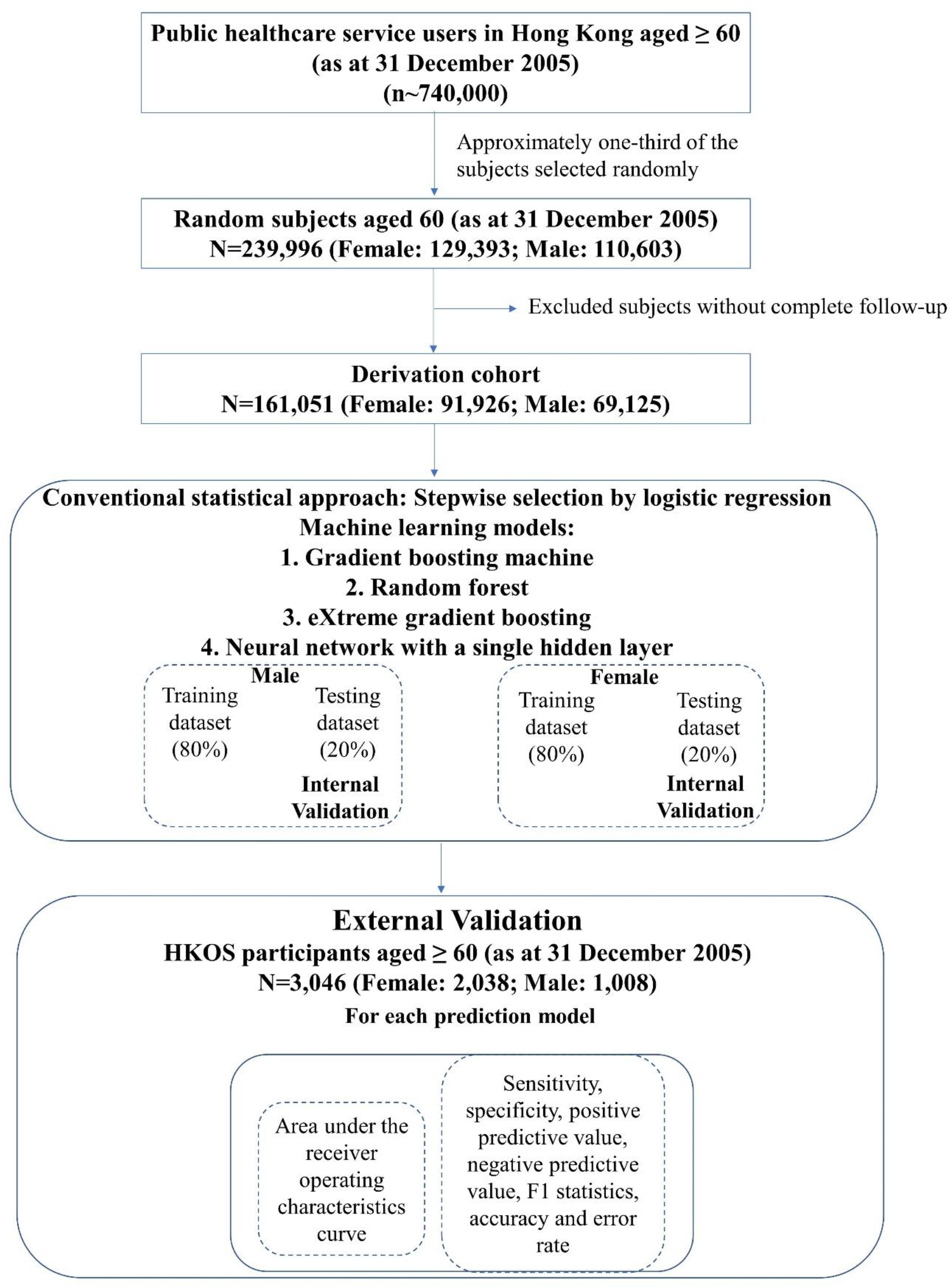
Study design and workflow of cohort derivation.

### Predictor variables

Potential predictors, including age on index date, all diagnosis and drug prescription records within one year preceding the index date, were retrieved from CDARS for individuals in the derivation and external validation cohorts. The presence or absence of each diagnosis code (as sub-chapters of ICD-9-CM) was recorded as binary coding using the icd package^14^ in R. Whether an individual was prescribed a class of drug (as BNF codes including chapters and sections) was also recorded as binary coding. Out of 395 potential predictors, 163 diagnosis and drug prescription variables with zero or near-zero variance (binary variables with ≤0.1% prevalence in the sex-stratified cohort) were excluded, leaving 232 potential predictor variables to train the prediction models.

### Development of prediction models

For the conventional statistical model, all potential predictors were included at the start, followed by a stepwise selection by logistic regression (LR) which added and dropped predictors to identify a model with the lowest Akaike Information Criteria (AIC),^15^ penalizing addition of variables into the model. An R package, “MASS”, was employed to implement the stepwise algorithm for LR.^16^ Four ML algorithms (including gradient boosting machine [GBM], random forest [RF], eXtreme gradient boosting [xgbTree], and neural networks with a single hidden layer [nnet]) were adopted to train the prediction model, utilizing the caret package in R.^17^ For each algorithm, hyperparameters were optimized with 10 repeats of 10-fold cross-validation to maximize the area under the receiver operating characteristic (ROC) curve (AUC) of the training model.

### Evaluation of prediction models

The general diagnostic accuracy of each model was evaluated by the AUC in the internal testing and external validation datasets. The optimal cut-off value for hip fracture risk classification was determined based on the ROC analysis of the training dataset using the Youden’s index.^18^ The sensitivity, specificity, PPV, negative predictive value (NPV), F1 statistics, accuracy and error rate were evaluated for each prediction model in the internal testing and external validation cohorts. DeLong’s test was used to compare the AUC of two models. With the LR model as reference, whether the ML algorithms had improvement in discrimination performance were assessed using the category-less net reclassification index (NRI) and integrated discrimination improvement index (IDI), which were computed using the Hmisc package^19^ in R. As a measure of both discrimination and calibration,^20^ the Brier score was calculated as the mean squared error between the actual event (fracture) and estimated probability.^21^ The calibration slope, intercept, and the Spiegelhalter Z-test (with perfect calibration as the null hypothesis)^22^ were computed using the rms package^23^ in R. Smaller Brier score, insignificant Spiegelhalter Z-test, a calibration slope closer to 1 and intercept closer to 0 imply better calibration. The observed and predicted probability of different models in external validation were presented as calibration curves.

### Ethics

The study protocol was approved by the institutional review board of the University of Hong Kong and the HA Hong Kong West Cluster (reference: UW 19-798), and the Hong Kong Polytechnic University (reference: HSEARS20201109004).

## Results

### Cohort participants

Figure 1 outlines the workflow in selecting study subjects of the derivation cohorts. The derivation cohort comprised 161,051 individuals (91,926 female; 69,125 male). Their baseline characteristics are presented in Table 1. The proportion of hip fracture cases in the derivation cohort was preserved in the constituting training and testing cohorts. In the female derivation cohort, 10.3% of the subjects had hip fracture within the 10-year follow-up (Table 1a). Only 6% subjects in the male derivation cohort had hip fracture events within the follow-up period (Table 1b). The baseline characteristics within one year prior to index date were similar among individuals in the training and internal testing cohorts (Table 1). Compared to the derivation cohort, individuals in the external validation cohort were younger and had fewer hip fracture cases during follow-up (Table 1). Some known risk factors of fracture, such as diagnosis of cardiovascular disease (CVD), prescription records of drugs for rheumatic diseases and gout, and corticosteroids, were less prevalent in the external validation cohort.

**Table 1.** Characteristics of the cohort participants in primary analysis.

(a) Characteristics of female cohort participants in the prediction model of 10-year risk of hip fracture.
| Characteristics | Derivation Cohort |  | External Validation Cohort<br>n=2,038 |
| --- | --- | --- | --- |
|  | Training Cohort<br>n=73,541 | Testing Cohort<br>n=18,385 |  |
| **Hip fracture cases within 10-year follow-up\*** | 7568 (10.3) | 1892 (10.3) | 145 (7.1)^ |
| **Age on index date** | 71.64 ± 7.42 | 71.58 ± 7.40 | 70.22 ± 6.87^ |
| *Diagnosis (1-year prior to index date)* |  |  |  |
| **Chronic obstructive pulmonary disease and allied conditions** | 574 (0.8) | 130 (0.7) | 10 (0.5) |
| **Any cancer** |  |  |  |
| **Malignant Neoplasm Of Lip, Oral Cavity, And Pharynx** | 15 (0.0) | 4 (0.0) | 0 (0.0) |
| **Malignant Neoplasm Of Digestive Organs And Peritoneum** | 140 (0.2) | 40 (0.2) | 2 (0.1) |
| **Malignant Neoplasm Of Respiratory And Intrathoracic Organs** | 22 (0.0) | 3 (0.0) | 0 (0.0) |
| **Malignant Neoplasm Of Bone, Connective Tissue, Skin, And Breast** | 135 (0.2) | 36 (0.2) | 6 (0.3) |
| **Malignant Neoplasm Of Genitourinary Organs** | 98 (0.1) | 30 (0.2) | 2 (0.1) |
| **Malignant Neoplasm Of Other And Unspecified Sites** | 55 (0.1) | 20 (0.1) | 1 (0.0) |
| **Malignant Neoplasm Of Lymphatic And Hematopoietic Tissue** | 18 (0.0) | 5 (0.0) | 3 (0.1)^ |
| **Cardiovascular disease** |  |  |  |
| **Chronic Rheumatic Heart Disease** | 64 (0.1) | 23 (0.1) | 1 (0.0) |
| **Hypertensive Disease** | 1937 (2.6) | 497 (2.7) | 48 (2.4) |
| **Ischemic Heart Disease** | 780 (1.1) | 230 (1.3)# | 17 (0.8) |
| **Diseases Of Pulmonary Circulation** | 18 (0.0) | 3 (0.0) | 0 (0.0) |
| **Other Forms Of Heart Disease** | 952 (1.3) | 232 (1.3) | 15 (0.7)^ |
| **Cerebrovascular Disease** | 829 (1.1) | 217 (1.2) | 12 (0.6)^ |
| **Diseases Of Arteries, Arterioles, And Capillaries** | 130 (0.2) | 25 (0.1) | 4 (0.2) |
| **Diseases Of Veins And Lymphatics, And Other Diseases Of Circulatory System** | 383 (0.5) | 97 (0.5) | 13 (0.6) |
| **Psychotic conditions, including dementias, alcohol-induced mental disorders** | 374 (0.5) | 101 (0.5) | 4 (0.2) |
| **Rheumatism, Excluding The Back** | 296 (0.4) | 86 (0.5) | 11 (0.5) |
| **Nephritis, Nephrotic Syndrome, And Nephrosis** | 170 (0.2) | 44 (0.2) | 2 (0.1) |
| **Diseases of Endocrine Glands, including diabetes mellitus, disorders of pituitary and parathyroid glands, ovarian dysfunction** | 1290 (1.8) | 353 (1.9) | 25 (1.2) |
| **Previous fracture** |  |  |  |
| **Fracture Of Skull** | 23 (0.0) | 9 (0.0) | 2 (0.1) |
| **Fracture Of Neck And Trunk** | 188 (0.3) | 54 (0.3) | 6 (0.3) |
| **Fracture Of Upper Limb** | 496 (0.7) | 123 (0.7) | 15 (0.7) |
| **Fracture Of Lower Limb** | 599 (0.8) | 144 (0.8) | 23 (1.1) |
| *Drug Prescription (within 1-year prior to index date)* |  |  |  |
| **Any antidepressants (BNF 4.3)** | 2856 (3.9) | 764 (4.2) | 62 (3.0)^ |
| **Drugs used in rheumatic diseases and gout (BNF 10.1)** | 13516 (18.4) | 3380 (18.4) | 270 (13.2)^ |
| **Corticosteroids** |  |  |  |
| **Respiratory (BNF 3.2)** | 1294 (1.8) | 281 (1.5)# | 27 (1.3) |
| **Endocrine (BNF 6.3)** | 1875 (2.5) | 484 (2.6) | 25 (1.2)^ |
| **Topical (BNF 13.4)** | 10165 (13.8) | 2454 (13.3) | 175 (8.6)^ |

| Characteristics | Derivation Cohort |  | External Validation Cohort<br>n=1,008 |
| --- | --- | --- | --- |
|  | Training Cohort<br>n=55,301 | Testing Cohort<br>n=13,824 |  |
| **Hip fracture cases within 10-year follow-up\*** | 3301 (6.0) | 825 (6.0) | 36 (3.6)^ |
| **Age on index date** | 70.02 ± 6.58 | 70.02 ± 6.55 | 69.58 ± 5.8^ |
| *Diagnosis (within 1-year prior to index date)* |  |  |  |
| **Chronic obstructive pulmonary disease and allied conditions** | 716 (1.3) | 174 (1.3) | 8 (0.8) |
| **Any cancer** |  |  |  |
| **Malignant Neoplasm Of Lip, Oral Cavity, And Pharynx** | 31 (0.1) | 4 (0.0) | 1 (0.1) |
| **Malignant Neoplasm Of Digestive Organs And Peritoneum** | 186 (0.3) | 46 (0.3) | 3 (0.3) |
| **Malignant Neoplasm Of Respiratory And Intrathoracic Organs** | 62 (0.1) | 15 (0.1) | 0 (0.0) |
| **Malignant Neoplasm Of Bone, Connective Tissue, Skin, And Breast** | 21 (0.0) | 6 (0.0) | 0 (0.0)^ |
| **Malignant Neoplasm Of Genitourinary Organs** | 217 (0.4) | 66 (0.5) | 2 (0.2) |
| **Malignant Neoplasm Of Other And Unspecified Sites** | 69 (0.1) | 18 (0.1) | 1 (0.1) |
| **Malignant Neoplasm Of Lymphatic And Hematopoietic Tissue** | 26 (0.0) | 9 (0.1) | 0 (0.0) |
| **Cardiovascular disease** |  |  |  |
| **Chronic Rheumatic Heart Disease** | 42 (0.1) | 11 (0.1) | 1 (0.1) |
| **Hypertensive Disease** | 1439 (2.6) | 374 (2.7) | 15 (1.5)^ |
| **Ischemic Heart Disease** | 990 (1.8) | 240 (1.7) | 7 (0.7)^ |
| **Diseases Of Pulmonary Circulation** | 11 (0.0) | 7 (0.1) | 1 (0.1) |
| **Other Forms Of Heart Disease** | 756 (1.4) | 211 (1.5) | 9 (0.9) |
| **Cerebrovascular Disease** | 850 (1.5) | 196 (1.4) | 10 (1.0) |
| **Diseases Of Arteries, Arterioles, And Capillaries** | 151 (0.3) | 27 (0.2) | 2 (0.2) |
| **Diseases Of Veins And Lymphatics, And Other Diseases Of Circulatory System** | 380 (0.7) | 108 (0.8) | 6 (0.6) |
| **Psychotic conditions, including dementias, alcohol-induced mental disorders** | 158 (0.3) | 43 (0.3) | 1 (0.1) |
| **Rheumatism, Excluding The Back** | 183 (0.3) | 46 (0.3) | 3 (0.3) |
| **Nephritis, Nephrotic Syndrome, And Nephrosis** | 170 (0.3) | 61 (0.4)# | 3 (0.3) |
| **Diseases of Endocrine Glands, including diabetes mellitus, disorders of pituitary and parathyroid glands, ovarian dysfunction** | 967 (1.7) | 229 (1.7) | 7 (0.7)^ |
| **Previous fracture** |  |  |  |
| **Fracture Of Skull** | 20 (0.0) | 6 (0.0) | 0 (0.0) |
| **Fracture Of Neck And Trunk** | 68 (0.1) | 18 (0.1) | 0 (0.0) |
| **Fracture Of Upper Limb** | 117 (0.2) | 42 (0.3) | 2 (0.2) |
| **Fracture Of Lower Limb** | 209 (0.4) | 59 (0.4) | 2 (0.2) |
| *Drug Prescription (within 1-year prior to index date)* |  |  |  |
| **Any antidepressants (BNF 4.3)** | 1117 (2.0) | 248 (1.8) | 15 (1.5) |
| **Drugs used in rheumatic diseases and gout (BNF 10.1)** | 9908 (17.9) | 2511 (18.2) | 124 (12.3)^ |
| **Corticosteroids** |  |  |  |
| **Respiratory (BNF 3.2)** | 1346 (2.4) | 307 (2.2) | 13 (1.3)^ |
| **Endocrine (BNF 6.3)** | 1863 (3.4) | 461 (3.3) | 19 (1.9)^ |
| **Topical (BNF 13.4)** | 7175 (13.0) | 1834 (13.3) | 95 (9.4)^ |

For continuous variables, data are presented as mean ± standard deviation. Continuous variables (with normal distribution) between groups were compared using t-test, while those with non-normal distribution were compared using Kruskal-Wallis test. For binary variables, data are presented as numbers (percentage), comparison between groups was done using chi-square test.
\* The overall proportion of hip fracture cases were preserved in the 80% training and 20% testing cohorts.

### Performance of prediction models

The discrimination performance metrics of the female prediction models in internal and external validation cohorts are presented in Table 2. In the internal validation cohort, the stepwise selection by LR, GBM and xgbTree models attained the highest AUC of 0.815 (95% Confidence Interval: 0.805-0.825). Using the Youden’s index to determine the optimal threshold for hip fracture classification, the LR model had moderate sensitivity and specificity (>0.7) (Table 2 and Supplementary Figure S1). All the ML algorithms had statistically significant and negative IDI and NRI with reference to the LR model, implying that they could not further improve the discrimination performance (Supplementary Table S1). The DeLong’ test showed that the AUC of the LR model was significantly higher than the RF and nnet models (Table 2). The LR model was well-calibrated, as suggested by the small Brier’s score and insignificant Spiegelhalter Z-test (Supplementary Table S2 and Supplementary Figure S2). In external validation, the LR model attained a high AUC of 0.841 (0.807-0.87). With the threshold defined by the Youden’s index, the LR model also had moderate sensitivity (0.69) but high specificity (0.82). Its AUC was significantly higher than the RF model, but comparable to other ML models with AUC in the range of 0.832-0.845 (Table 2 and Supplementary Figure S3). The negative IDI and NRI showed that the ML models could not further improve the discrimination performance of the LR model (Supplementary Table S1). The LR model also had adequate calibration in external validation (Supplementary Table S2 and Supplementary Figure S4).

**Table 2.** Discrimination performance of hip fracture risk prediction models for female.

| Algorithm used in model development | Stepwise selection by logistic regression | Gradient boosting machine | Random forest | eXtreme gradient boosting | Neural networks with a single hidden layer |
| --- | --- | --- | --- | --- | --- |
| **Derivation cohort** |  |  |  |  |  |
| *Training cohort* |  |  |  |  |  |
| **AUC (95% CI)** | 0.823 (0.818-0.827) | 0.823 (0.818-0.828) | 0.996 (0.996-0.997) | 0.826 (0.821-0.831) | 0.825 (0.82-0.83) |
| *Testing cohort* |  |  |  |  |  |
| **AUC (95% CI)** | 0.815 (0.805-0.825) | 0.815 (0.805-0.825) | 0.78 (0.769-0.791) | 0.815 (0.805-0.825) | 0.803 (0.792-0.813) |
| **Sensitivity** | 0.721 | 0.754 | 0.5 | 0.757 | 0.724 |
| **Specificity** | 0.754 | 0.724 | 0.868 | 0.721 | 0.739 |
| **PPV** | 0.252 | 0.239 | 0.302 | 0.237 | 0.241 |
| **NPV** | 0.959 | 0.962 | 0.938 | 0.963 | 0.959 |
| **F1** | 0.373 | 0.362 | 0.376 | 0.361 | 0.362 |
| **Accuracy** | 0.751 | 0.727 | 0.83 | 0.724 | 0.737 |
| **Error** | 0.249 | 0.273 | 0.17 | 0.276 | 0.263 |
| **Delong's test p-value** | Reference | 0.95 | <0.001 | 0.975 | <0.001 |
| **External validation cohort** |  |  |  |  |  |
| **AUC (95% CI)** | 0.841 (0.807-0.87) | 0.845 (0.811-0.879) | 0.813 (0.779-0.848) | 0.842 (0.808-0.877) | 0.832 (0.797-0.867) |
| **Sensitivity** | 0.69 | 0.724 | 0.51 | 0.731 | 0.731 |
| **Specificity** | 0.817 | 0.802 | 0.895 | 0.797 | 0.793 |
| **PPV** | 0.224 | 0.219 | 0.271 | 0.216 | 0.213 |
| **NPV** | 0.972 | 0.974 | 0.96 | 0.975 | 0.975 |
| **F1** | 0.338 | 0.336 | 0.354 | 0.333 | 0.33 |
| **Accuracy** | 0.808 | 0.796 | 0.868 | 0.792 | 0.789 |
| **Error** | 0.192 | 0.204 | 0.133 | 0.208 | 0.211 |
| **Delong's test p-value** | Reference | 0.149 | 0.016 | 0.687 | 0.291 |

The discrimination performance of the prediction models developed for male are presented in Table 3. In internal validation, although the xgbTree model had a significantly higher AUC of 0.825 (0.809-0.84) than the LR model (0.818 [0.801-0.834]) (Table 3, Supplementary Figure S5), the discrimination performance of LR model outperformed other models as indicated by the negative IDI and NRI of the ML models (Supplementary Table S3). Adequate calibration was also observed for the LR model (Supplementary Table S4 and Supplementary Figure S6). In external validation, the LR model had a high AUC of 0.898 (0.857-0.939), which was significantly higher than the RF model, but comparable to other ML models with AUC in the range of 0.898-0.905 (Table 3, Supplementary Figure S7). The IDI and NRI of the GBM, RF and xgbTree models were statistically significant and negative, implying that they could not improve the discrimination performance of the LR model (Supplementary Table S3). The negative IDI of the nnet model reached statistical significance, but not the NRI (Supplementary Table S3). Moreover, the calibration was inadequate in external validation for all the male prediction models (Supplementary Table S4 and Supplementary Figure S8).

**Table 3.** Discrimination performance of hip fracture risk prediction models for male.

| Algorithm used in model development | Stepwise selection by logistic regression | Gradient boosting machine | Random forest | eXtreme gradient boosting | Neural networks with a single hidden layer |
| --- | --- | --- | --- | --- | --- |
| **Derivation Cohort** |  |  |  |  |  |
| *Training cohort* |  |  |  |  |  |
| **AUC (95% CI)** | 0.826 (0.819-0.834) | 0.825 (0.818-0.833) | 0.996 (0.995-0.997) | 0.834 (0.827-0.841) | 0.826 (0.819-0.834) |
| *Testing cohort* |  |  |  |  |  |
| **AUC (95% CI)** | 0.818 (0.801-0.834) | 0.824 (0.808-0.839) | 0.775 (0.757-0.793) | 0.825 (0.809-0.84) | 0.818 (0.802-0.833) |
| **Sensitivity** | 0.744 | 0.742 | 0.416 | 0.736 | 0.727 |
| **Specificity** | 0.749 | 0.753 | 0.923 | 0.758 | 0.761 |
| **PPV** | 0.158 | 0.16 | 0.254 | 0.162 | 0.162 |
| **NPV** | 0.979 | 0.979 | 0.961 | 0.978 | 0.978 |
| **F1** | 0.261 | 0.263 | 0.315 | 0.265 | 0.264 |
| **Accuracy** | 0.749 | 0.752 | 0.892 | 0.756 | 0.759 |
| **Error** | 0.251 | 0.248 | 0.108 | 0.244 | 0.241 |
| **Delong's test p-value** | Reference | 0.066 | <0.001 | 0.019 | 0.878 |
| **External validation cohort** |  |  |  |  |  |
| **AUC (95% CI)** | 0.898 (0.857-0.939) | 0.898 (0.857-0.939) | 0.84 (0.783-0.896) | 0.9 (0.861-0.939) | 0.905 (0.863-0.947) |
| **Sensitivity** | 0.806 | 0.75 | 0.25 | 0.75 | 0.806 |
| **Specificity** | 0.817 | 0.81 | 0.957 | 0.824 | 0.823 |
| **PPV** | 0.14 | 0.127 | 0.176 | 0.136 | 0.144 |
| **NPV** | 0.991 | 0.989 | 0.972 | 0.989 | 0.991 |
| **F1** | 0.239 | 0.218 | 0.207 | 0.231 | 0.245 |
| **Accuracy** | 0.817 | 0.808 | 0.932 | 0.821 | 0.822 |
| **Error** | 0.184 | 0.193 | 0.069 | 0.179 | 0.178 |
| **Delong's test p-value** | Reference | 0.993 | 0.005 | 0.755 | 0.34 |

### Association of predictors with hip fracture

Since the discrimination performance of the LR model outperformed the ML models in both female and male in internal testing and external validation, the 20 top predictors adopted by the model having the strongest association with hip fracture are listed in Table 4. Eleven of them were among the top 20 in both the female and male prediction models.

**Table 4.** The top 20 predictors selected by stepwise selection by logistic regression models with the strongest association with hip fracture.

| Predictors | Training cohort |  |  |
| --- | --- | --- | --- |
|  | OR | (95% CI) | p-value |
| **Age on index date** | 1.161 | (1.156 - 1.165) | <0.001 |
| *Diagnosis* |  |  |  |
| Accidental falls | 1.673 | (1.403 - 1.996) | <0.001 |
| **Chronic obstructive pulmonary disease and allied Conditions** | 1.506 | (1.181 - 1.921) | <0.001 |
| Dorsopathies | 1.415 | (1.185 - 1.689) | <0.001 |
| Nephritis, nephrotic syndrome, and nephrosis | 2.299 | (1.548 - 3.417) | <0.001 |
| **Organic psychotic conditions** | 1.651 | (1.278 - 2.133) | <0.001 |
| *Drug prescription* |  |  |  |
| **Anaemias and some other blood disorders** | 1.52 | (1.293 - 1.786) | <0.001 |
| Antidepressant drugs | 1.231 | (1.082 - 1.402) | <0.001 |
| **Antiplatelet drugs** | 1.187 | (1.099 - 1.282) | <0.001 |
| **Beta-adrenoceptor blocking drugs** | 0.899 | (0.844 - 0.958) | <0.001 |
| **Bronchodilators** | 1.305 | (1.174 - 1.452) | <0.001 |
| **Drugs acting on the oropharynx** | 0.793 | (0.729 - 0.863) | <0.001 |
| **Drugs used in diabetes** | 1.753 | (1.638 - 1.875) | <0.001 |
| **Drugs used in parkinsonism and related disorders** | 2.255 | (1.869 - 2.721) | <0.001 |
| **Drugs used in psychoses and related disorders** | 1.369 | (1.153 - 1.626) | <0.001 |
| **Laxatives** | 1.275 | (1.186 - 1.37) | <0.001 |
| Miscellaneous drugs (Nutrition and blood) | 5.967 | (2.432-14.638) | <0.001 |
| Minerals | 1.161 | (1.055 - 1.278) | 0.002 |
| Positive inotropic drugs | 1.504 | (1.224 - 1.848) | <0.001 |
| **Vitamins** | 1.132 | (1.051 - 1.22) | 0.001 |

OR: Odds Ratio; CI: Confidence Interval. Predictors in bold were among the top 20 predictors in both female and male models.

| Predictors | Training cohort |  |  |
| --- | --- | --- | --- |
|  | OR | (95% CI) | p-value |
| **Age on index date** | 1.164 | (1.157 - 1.17) | <0.001 |
| *Diagnosis* |  |  |  |
| **Chronic obstructive pulmonary disease and allied conditions** | 1.63 | (1.243 - 2.138) | <0.001 |
| **Organic psychotic conditions** | 2.153 | (1.415 - 3.277) | <0.001 |
| Other accidents | 1.875 | (1.315 - 2.673) | <0.001 |
| Poisoning by drugs, medicinal and biological substances | 10.166 | (3.283 - 31.463) | <0.001 |
| *Drug prescription* |  |  |  |
| **Anaemias and some other blood disorders** | 1.608 | (1.266 - 2.042) | <0.001 |
| Antiepileptic drugs | 1.647 | (1.277 - 2.124) | <0.001 |
| **Antiplatelet drugs** | 1.437 | (1.295 - 1.594) | <0.001 |
| **Beta-adrenoceptor blocking drugs** | 0.841 | (0.763 - 0.926) | <0.001 |
| **Bronchodilators** | 1.547 | (1.359 - 1.761) | <0.001 |
| **Drugs acting on the oropharynx** | 0.804 | (0.708 - 0.913) | <0.001 |
| **Drugs used in diabetes** | 1.431 | (1.288 - 1.59) | <0.001 |
| **Drugs used in parkinsonism and related disorders** | 3.209 | (2.496 - 4.13) | <0.001 |
| **Drugs used in psychoses and related disorders** | 1.998 | (1.552 - 2.571) | <0.001 |
| Emollient and barrier preparations | 1.448 | (1.301 - 1.611) | <0.001 |
| Fluids and electrolytes | 1.307 | (1.134 - 1.507) | <0.001 |
| **Laxatives** | 1.373 | (1.237 - 1.524) | <0.001 |
| Lipid-regulating drugs | 0.744 | (0.648 - 0.855) | <0.001 |
| Miscellaneous drugs (skin) | 5.366 | (1.981 - 14.52) | <0.001 |
| **Vitamins** | 1.399 | (1.245 - 1.573) | <0.001 |

OR: Odds Ratio; CI: Confidence Interval. Predictors in bold were among the top 20 predictors in both female and male models.

## Discussion

In the current study, we utilized EHR of >160,000 individuals from a population-based cohort to develop 10-year sex-specific hip fracture risk prediction models in Hong Kong, using both conventional statistical approach and ML algorithms. The prediction models were validated in the internal testing cohort of public healthcare service users, and the external validation cohort of community-dwelling individuals. The conventional LR model outperformed the ML models in both female and male. In particular, the LR model for female was adequately calibrated, suggesting the potential usefulness clinically. To our knowledge, this is one of the largest samples used to develop hip fracture prediction models among the Asians.

One noticeable feature of our prediction models is that we included age, all diagnosis and drug prescription records from the electronic medical database as potential predictors, irrespective of their prior association with hip fracture. Most importantly, BMD data was not used in model development. Since the EHR was input by clinicians and healthcare professionals at patient visit, the readily available data enhanced the feasibility of integrating the prediction models into the routine clinical workflow of public healthcare setting in Hong Kong. Even in the absence of BMD data, the LR model for female had AUC>0.8 in both internal testing and external validation. In addition to adequate calibration, this model is likely to be clinically useful in risk stratification.^24^ Although the AUC of the LR model for male was also high in internal and external validation (>0.8), the external validation was inadequately calibrated, which may be attributed to the relatively small sample number of male participants in HKOS. Further validation of the male prediction models in independent cohort of larger sample size is warranted to evaluate its potential usefulness in hip fracture risk prediction. In comparison with existing fracture prediction tools, such as QFracture,^25^ FRAX^6^ and Garvan,^26^ they included only a pre-defined set of conventional risk factors of hip fracture in development of the prediction model. Notably, clinical parameters such as weight and/or height were used as the conventional predictors in FRAX^6^ and Garvan,^26^ if BMD data was unavailable. Conversely, our prediction models did not include any clinical parameters (such as weight, height, and BMD) as predictor. In addition, while the internal testing cohort consisted of public healthcare service users, our external validation cohort comprised the HKOS participants who were community-dwelling individuals, demonstrating the potentially high generalizability of our prediction models.

Several studies have adopted the ML approach to predict future fracture risk.^8-10^ A study utilized the national Danish patient data of 6,600 individuals to develop a 5-year hip fracture prediction model. With DXA data and laboratory tests, their prediction models had a good performance with AUC>0.9.^10^ Nevertheless, DXA screening is not easily accessible,^4,5^ limiting its generalizability. Another study used data of 5,130 individuals from the Osteoporosis Fractures in Men (MrOS) for predicting the major osteoporotic fracture. With the genetic risk score, BMD and other known risk factors as predictors, they developed a prediction model with AUC of 0.71.^8^ Since BMD, genotyping data and thus genetic risk score are not readily available among the public, this model also has limited generalizability. Another study used the administrative claims data of 288,086 individuals in Germany to develop an osteoporotic hip fracture prediction model with 4-year follow-up. Age, sex, history of fracture and medications known to be related to bone health were adopted as the predictors, attaining an AUC of 0.65 to 0.7.^9^ Compared to these ML studies, our current study had sufficient sample size and the longest follow-up of 10 years. Notably, some of our ML models still had good discrimination performance (AUC>0.8) even in the absence of BMD data. One plausible reason is the inclusion of all diagnosis and drug prescription records as potential predictors, as some comorbidities and drug use also contribute to BMD variation. This aligns with a previous proposal by the developers of fracture risk evaluation model (FREM) that the optimal prediction model should include both common (with known small or modest effects on fracture risk) and rare (whose relationship with fracture risk is yet to be revealed) risk factors ^27^. The FREM utilized all the ICD-10 codes available from the Danish national register (n=2,495,339) and applied backward selection by LR to develop one-year sex-stratified prediction models of hip fracture, attaining AUC of 0.87 and 0.85 for female and male respectively.^27^ The inclusion of drug prescription records in our models may contribute to the good discrimination performance despite the smaller sample size. More importantly, the best-performing models for both female and male in the current study were the stepwise selection by LR models, but not the ML models. This is in line with a systematic review reporting that ML algorithms did not necessarily have better performance than LR model in clinical risk prediction, despite the flexibility of including nonlinear association and interaction terms in the model.^28^

A number of conventional risk factors were selected by the LR models (Table 4), such as age,^29^ diagnosis and/or prescription records of accidental falls,^30^ CVD,^31^ chronic obstructive pulmonary diseases,^25^ Parkinson’s diseases,^32^ epilepsy,^33^ depression,^25^ diabetes,^34^ psychoses,^35^ and nutritional deficiencies.^36^ More importantly, our approach enables the identification of some relatively novel predictors of hip fracture. An example is drug prescription for anaemia and blood disorders, which was associated with higher odds of hip fracture (Table 4). This is consistent with our recent Mendelian randomization study that genetically determined red blood cell traits had positive causal effects on BMD.^37^ Individuals with blood disorders, such as anaemia, may have lifelong risk of osteoporosis and fracture. In general, vitamins, laxatives, emollient are prescribed for poor appetite, constipation, and dry skin respectively. Together with anaemia, they are signs of ageing or frailty, which are the most important risk factor for fracture. Nevertheless, the exact underlying mechanisms of how the novel predictors might influence bone health or hip fracture warrant future investigations. On the other hand, some predictors were sex-specific, probably attributed to their different prevalence between sexes. An example is the diagnosis of nephritis, nephrotic syndrome and nephrosis, which was included in the female prediction model (Table 4a). While chronic renal disease was adopted by QFracture as a risk factor irrespective of sex,^25^ its related diagnosis was identified as a female-specific risk factor in our study, which partially aligned with previous literature that hip fracture incidence among women with chronic kidney diseases was twice as high as that in men.^38^

This study has several strengths and may be clinically important. We developed sex-specific hip fracture prediction models without utilizing clinical measurement data, such as BMD and body mass index (BMI). Yet, the best-performing prediction models have good discrimination performance with AUC>0.8. The female model also has adequate calibration. Using EHR data as the only predictors enables the integration of the prediction models into routine clinical workflow in the public healthcare setting. Amid the COVID-19 pandemic, healthcare services and resources were diverted to combat COVID-19 and its related comorbidities from chronic diseases like osteoporosis.^39^ Moreover, the prediction models were externally validated in a community-dwelling cohort. Taken together, despite the limited resources, the hip fracture prediction models may be applied at both public healthcare service setting and the public at population-level, aiding to triage individuals who are at high risk of hip fracture for prioritized DXA scan, and subsequent treatment initiation. Such measures are expected to facilitate early prevention, timely diagnosis and treatment of osteoporosis.

Our study also has limitations. First, diagnosis and prescription records within 1 year prior to the index date were retrieved in the current study. Yet, the diagnosis of chronic diseases might not be repeatedly coded in CDARS, explaining why the top 20 predictors were mainly drug prescription variables. Notably, medication use was recorded in CDARS upon prescription regardless of the onset of the disease. Thus, the inclusion of drug prescription variables is complementary to the use of diagnosis variables. Second, the electronic medical database did not capture risk factors related to lifestyle (such as alcohol consumption and smoking) and clinical measurement (such as BMI and weight). Nevertheless, these may be proxied by the diagnosis and drug prescription records available. Third, the generalizability of the model to other populations is unclear.

In conclusion, we have developed and validated sex-specific hip fracture prediction tools at population-level in Hong Kong using EHR. Notably, the good discrimination and calibration performance of the LR model for female was validated in both internal and external cohorts, implying that the model may be clinically useful and generalizable to the public. Despite the high discrimination performance, the LR model for male would require additional calibration in independent cohorts. By using EHR as predictors, it is expected that the prediction model could be integrated to the routine clinical workflow, assisting clinicians to identify people who are at high risk of hip fracture for DXA scan. These measures may facilitate early prevention, timely diagnosis and treatment of osteoporosis.

## Contributors

GHL and CLC contributed to conceptualisation and study design, and act as guarantors for the study. GHL performed statistical analysis, implemented development and validation of prediction models, and drafted the manuscript. CLC, KCT and AWK contributed to the data resources. CLC, KCT, AWK, TCK, WCL, JSW, WWH, CF and ICW provided critical input to the analyses and discussion. All authors contributed to the data interpretation, critically reviewed and revised the manuscript, and approved the final manuscript.

## Data Availability

This study is conducted based on the anonymised dataset from the CDARS. We are unable to share the CDARS data used in this study since the data custodian, the Hong Kong Hospital Authority, has not provided us the permission. Nevertheless, CDARS data can be accessed via the Hospital Authority Data Sharing Portal for research purpose (https://www3.ha.org.hk/data).

